# Short Report. Intersectional Inequalities in Mental Health Need and Youth Justice Contact among Pupils enrolled in Alternative Provision

**DOI:** 10.64898/2026.11.08.26367542

**Authors:** Sorcha Ní Chobhthaigh, Camille Cox, Twyla Greenway-Bailey, Josephine Musanu, Mel Green, Amie Buhari, Mina Mawi, Cyra Neave, Rochelle A. Burgess, Matthew A. Jay, Delan Devakumar

## Abstract

**Background:** Unmet mental health and education needs increase the risk of youth justice contact. Understanding inequalities in alternative provision (AP) and how these relate to school recorded mental health needs or police criminalisation may identify opportunities to intervene.

**Methods:** We analysed population-level linked school and police administrative data for 3.7m pupils (born 1997-2003) who attended state schools in England. We examined differences by racial-ethnic group and gender in rates and timing of AP enrolment and recording patterns of mental health-related provision and youth justice contacts among pupils with AP enrolment.

**Results:** Our findings indicate heightened mental health need and vulnerability to criminalisation among pupils recorded with AP enrolment. Enrolment was highest among Irish Traveller and Romani pupils, followed by Black Caribbean and Mixed White–Black Caribbean pupils. Boys were enrolled more than girls, but some girls had higher rates than boys from other groups. Over 50% boys from Black Caribbean and Mixed White–Black Caribbean backgrounds with AP enrolment had a youth justice contact, the majority of whom had recorded mental health needs.

**Conclusion:** Heightened need and intersectional inequalities among pupils enrolled in AP highlight the importance of a co-ordinated public health approach to understanding and responding to children’s needs.

## Introduction

Alternative provision (AP) in England refers to education outside of mainstream or special schools for pupils who ‘would not otherwise receive suitable education’ (1). While attendance may be due to illness, pregnancy or other types of Special Educational Needs and Disability (SEND), pupil pathways to AP are predominantly related to experiences of exclusions or being ‘managed’ out of mainstream schools, followed by specified Social, Emotional and Mental Health (SEMH) needs (2,3). Qualitative reports highlight that these pupils’ complex mental health needs remain inadequately supported in AP (4,5), risking compounded unmet need and increased risk of youth justice contacts (6).

Inequalities based on racial-ethnic group and gender have been documented among pupils enrolled in AP (7), aligning with inequities in exclusions, referrals to mental health care and policing practices (8–10). A gap remains in understanding inequalities in AP at the intersection of both racial-ethnic group and gender, and how these relate to school identification of SEND-for-SEMH and police criminalisation of need.

## Methods

We analysed linked de-identified administrative data from the Police National Computer (PNC) and National Pupil Database (NPD). The PNC contains information on recordable offences (cautions and convictions) in England and Wales. The NPD contains information about pupils in state-maintained education in England. We included all pupils born 1997-2003 who attended state-school in England at any point during mandatory school years (age 5-16) (For more details see: (9)).

We examined outcomes at the intersection of racialisation and gender, using the NPD recording of “ethnicity” as a proxy for racialisation. Due to small numbers, we merged ‘Asian Other & Chinese’ groups (Supplementary Table 1). Pupils without “ethnicity” data (n=18,355) were excluded. Gender was recorded in NPD as male or female, representing assigned “sex”.

**Table 1:**
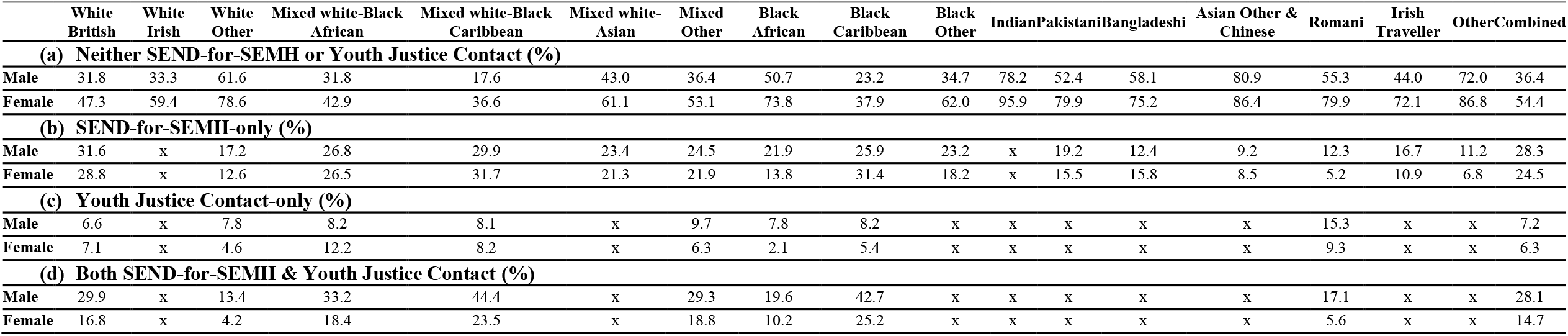
Recording Patterns: Proportions of (a) Neither SEND-for-SEMH or Youth Justice Contact (b) SEND-for-SEMH-only (c) Youth Justice Contact- only (d) Both Among Pupils Ever Recorded with AP Enrolment. In table ‘x’ used to denote suppression due to small cell counts.

|  | **White British** | **White Irish** | **White Other** | **Mixed white-Black African** | **Mixed white-Black Caribbean** | **Mixed white-Asian** | **Mixed Other** | **Black African** | **Black Caribbean** | **Black Other** | **Indian** | **Pakistani** | **Bangladeshi** | **Asian Other & Chinese** | **Romani** | **Irish Traveller** | **Other** | **Combined** |
|---|---|---|---|---|---|---|---|---|---|---|---|---|---|---|---|---|---|---|
| **(a) Neither SEND-for-SEMH or Youth Justice Contact (%)** |  |  |  |  |  |  |  |  |  |  |  |  |  |  |  |  |  |  |
| **Male** | 31.8 | 33.3 | 61.6 | 31.8 | 17.6 | 43.0 | 36.4 | 50.7 | 23.2 | 34.7 | 78.2 | 52.4 | 58.1 | 80.9 | 55.3 | 44.0 | 72.0 | 36.4 |
| **Female** | 47.3 | 59.4 | 78.6 | 42.9 | 36.6 | 61.1 | 53.1 | 73.8 | 37.9 | 62.0 | 95.9 | 79.9 | 75.2 | 86.4 | 79.9 | 72.1 | 86.8 | 54.4 |
| **(b) SEND-for-SEMH-only (%)** |  |  |  |  |  |  |  |  |  |  |  |  |  |  |  |  |  |  |
| **Male** | 31.6 | x | 17.2 | 26.8 | 29.9 | 23.4 | 24.5 | 21.9 | 25.9 | 23.2 | x | 19.2 | 12.4 | 9.2 | 12.3 | 16.7 | 11.2 | 28.3 |
| **Female** | 28.8 | x | 12.6 | 26.5 | 31.7 | 21.3 | 21.9 | 13.8 | 31.4 | 18.2 | x | 15.5 | 15.8 | 8.5 | 5.2 | 10.9 | 6.8 | 24.5 |
| **(c) Youth Justice Contact-only (%)** |  |  |  |  |  |  |  |  |  |  |  |  |  |  |  |  |  |  |
| **Male** | 6.6 | x | 7.8 | 8.2 | 8.1 | x | 9.7 | 7.8 | 8.2 | x | x | x | x | x | 15.3 | x | x | 7.2 |
| **Female** | 7.1 | x | 4.6 | 12.2 | 8.2 | x | 6.3 | 2.1 | 5.4 | x | x | x | x | x | 9.3 | x | x | 6.3 |
| **(d) Both SEND-for-SEMH & Youth Justice Contact (%)** |  |  |  |  |  |  |  |  |  |  |  |  |  |  |  |  |  |  |
| **Male** | 29.9 | x | 13.4 | 33.2 | 44.4 | x | 29.3 | 19.6 | 42.7 | x | x | x | x | x | 17.1 | x | x | 28.1 |
| **Female** | 16.8 | x | 4.2 | 18.4 | 23.5 | x | 18.8 | 10.2 | 25.2 | x | x | x | x | x | 5.6 | x | x | 14.7 |

NPD recording of AP enrolment indicates if a pupil was documented as enrolled in alternative provision or a pupil referral unit. ‘SEND-for-SEMH’ refers to any NPD record of SEND for Behaviour, Emotional & Social Difficulties up to 2013/2014 and Social, Emotional and Mental Health (SEMH) difficulties from 2014/2015 onwards. ‘Youth justice contact’ refers to any documented PNC record of a caution or conviction for a recordable offence between age 10-17.

We calculated (1) rates of AP enrolment, (2) median age-at-first recording using an approximate date of birth derived from NPD, and (3) among pupils with AP enrolment, the proportion recorded with SEND-for-SEMH only, youth justice contact only or both.

Data processing and analyses were completed with R 4.4.4 (11) in the Office for National Statistics Secure Research Service.

## Results

Our cohort included 3,694,590 pupils (Supplementary: Figure 1 & Table 2).

**Figure 1.**
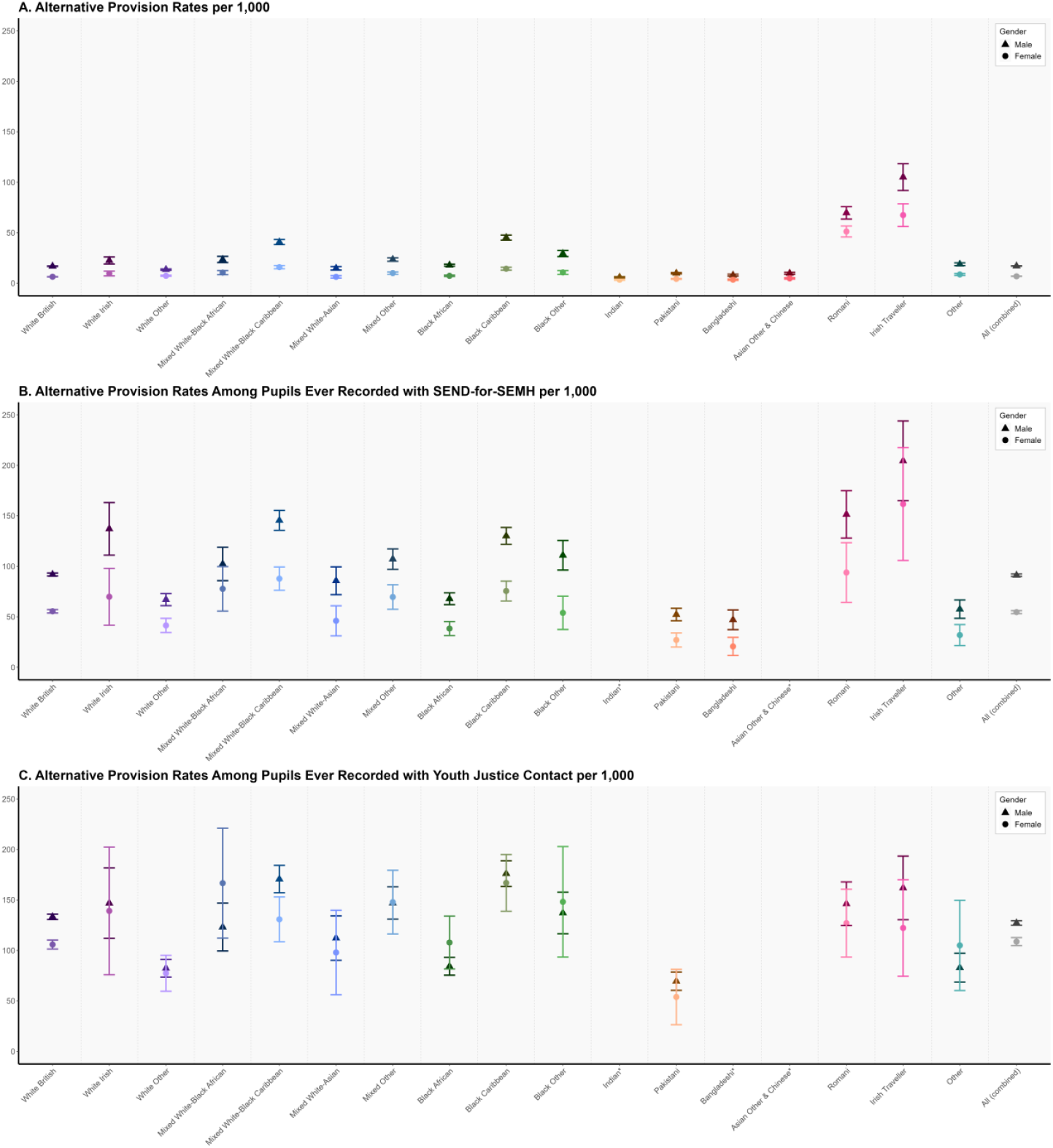
Rates of AP enrolment per 1,000 pupils (a) All pupils ever recorded with AP enrolment during their enrolment period (b) Among pupils ever recorded with SEND-for-SEMH (c) Among pupils ever recorded with Youth Justice Contact

### Rates

Rates of AP enrolment were highest among Romani and Irish Traveller pupils, followed by Black Caribbean and Mixed White-Black Caribbean pupils (Figure 1, Supplementary Table 3). Overall, boys were enrolled at higher rates, though rates for Irish Traveller and Romani girls were higher than boys from all other racial-ethnic groups.

AP rates were higher among pupils with youth justice contact (1.5-7.9 times higher boys; 1.8-16.6 times higher girls) and SEND-for-SEMH (1.9-6.1 times higher boys; 1.8-8.7 times higher girls).

### Age

For most groups, median age-at-first AP enrolment clustered at 13-14 years for boys and 14-15 for girls, younger for Irish Traveller pupils age 12 (Supplementary Table 4).

For pupils enrolled in AP, across groups the median age-at-first youth justice contact was 14.0 for boys and 14.1 for girls.

### Recording Patterns

Of pupils ever recorded with AP enrolment, 56% boys and 39% girls were recorded with SEND-for-SEMH, and 35% boys and 21% girls were recorded with youth justice contact, the majority of whom had recorded SEND-for-SEMH. Across racial-ethnic groups, recording patterns varied by approximately 60 percentage points within genders (Table 1); the proportion with neither record ranged from 17.6% (Mixed White-Black Caribbean boys) to 95.9% (Indian girls).

Where data was available, dual recordings were highest among Mixed White-Black Caribbean (boys: 44.4%; girls: 23.5%) and Black Caribbean (boys: 42.7%; girls: 25.2%) pupils.

## Discussion

### What is already known on this topic

Intersectional discrimination exists in recorded SEND-for-SEMH, use of exclusions and pathways to appropriate mental health care (8,12). Indian and Asian Other & Chinese pupils are least likely to receive SEND-for-SEMH following severe absences and exclusions. Black Caribbean, Mixed White- Black Caribbean, Romani and Irish Traveller pupils are more likely to be permanently excluded prior to/without receiving SEND-for-SEMH influencing their likelihood of unmet need and AP enrolment. At the same time, discrimination in policing practices (13,14) heightens the risk of criminalisation of need among racially and ethnically minoritised children. Insufficiently supported mental health needs and transition to secondary school leave children vulnerable to escalating need and pathways to criminalisation (15,16).

### What this study adds

Our findings highlight heightened mental health need and vulnerability to criminalisation among pupils recorded with AP enrolment. Pupils enrolled in AP with youth justice contact indicate unmet need and risk escalating cycles of need without sufficient therapeutic support. Substantial variation exists in enrolment and recording patterns at the intersection of racialisation and gender. Irish Traveller pupils experience the highest rates and youngest enrolment. For Black Caribbean and Mixed White-Black Caribbean, over 50% boys and approximately 30% girls enrolled in AP had a youth justice contact, the majority of whom had identified mental health needs. The timing of AP enrolment following the transition period indicates an opportunity for targeted intervention in mainstream schools to identify and address needs prior to escalation.

### Limitations

Small numbers prevent reporting of specific recording patterns for some groups, of note, for Romani and Irish Traveller pupils who have higher rates of AP enrolment, SEND-for-SEMH and youth justice contacts (9). The AP census began 2008/2009 as such AP enrolment is likely underestimated, and the Pupil Referral Unit census was only available 2010-2013. PNC does not capture interactions not resulting in caution/conviction, e.g., ‘stop and search’ or police involvement in school discipline - with documented inequities in these practices (13).

## Conclusion

Our findings indicate intersectional inequities in AP enrolment and recording patterns. Discrimination in responses to need in mainstream schools, manifesting as under-supported or over-punished need (8), drive trajectories towards AP. While AP has the potential to act as a site of therapeutic intervention that appropriately supports complex mental health needs and enables pupils to engage with education, wide variation and inconsistencies in provision and staff training is reported, risking escalating unmet need (4). Risks may be amplified for racially and ethnically minoritised pupils due to discrimination in the use of harmful disciplinary practices which collide with discrimination in policing (12,14). Without establishing a standard of therapeutic, evidence-based and trauma-informed AP alongside mandatory anti-discrimination training and accountability mechanisms, these failures increase the risk that instead of acting as a site of healing, AP serves as a ‘pipeline’ from mainstream school-to-prison.

## Ethics

This study uses linked, de-identified data from the National Public Database and Police National Computer as part of the Ministry of Justice (MoJ) and Administrative Data Research (ADR) UK funded data linkage program, Data First. Data access was granted by the data owners, MoJ and DfE, February 2024. Consent from individuals is not required. Study-specific ethical approval was obtained (10.03.2023) from UCL Research Ethics Committee (24643/001).

## Data availability

This study uses routinely collected administrative data from the Ministry of Justice (MoJ) and Department for Education (DfE). The data is not publicly available and can be accessed by researchers by applying directly to the MoJ and DfE.

## Author contributions

SNC is the guarantor: SNC had access to the data in the study and takes responsibility for the integrity of the data and the accuracy of the data analysis. SNC conceptualised and designed the study, performed the data analyses, and wrote the initial draft manuscript. CC, JM and TGB contributed to study design, analysis decisions, interpretation of results and co-development of recommendations. CN, MG, MM, and AB contributed to interpretation of results and co-development of recommendations. MJ supported data management, code development and supervised the analytical approach. RB supervised the participatory methodology. DD provided supervisory support throughout. All authors reviewed, commented, and approved the final manuscript.

## Acknowledgements

We would like to express our appreciation to additional and former stakeholder advisors (in addition to those who are named authors). Their meaningful contributions and thoughtful insights shaped this project, contextualisation and co-developed recommendations. We also extend our wholehearted gratitude to the No More Exclusions coalition for their valuable feedback and insights.

We gratefully acknowledge all children, young people and families whose de-identified data are used in this research.

This work was undertaken in the Office for National Statistics Secure Research Service using Data First linked administrative data. The use of the data in this work does not imply the endorsement of the ONS or data owners (e.g. MoJ and DfE) in relation to the interpretation or analyses of the statistical data. This work uses research datasets which may not exactly reproduce National Statistics aggregates. National statistics follow consistent statistical conventions over time and cannot be compared to Data First linked datasets.

The views in this publication do not necessarily reflect the views of UCL.

## Supplementary Files

- Supplementary Table 1: Racial-Ethnic Groups
- Supplementary Figure 1: Data Flow Summary
- Supplementary Table 2: Cohort Overview with Median Age at School Enrolment
- Supplementary Table 3: Rates per 1,000 children and young people
- Supplementary Table 4: Median Age-at-First

