## Supplementary Files for "Short Report. Intersectional Inequalities in Mental Health Need and Youth Justice Contact among Pupils enrolled in Alternative Provision"

**Supplementary Table 1: Racial-Ethnic Groups**

| ***National Pupil Database 'Ethnic Minor' variable categories*** | ***Study racial-ethnic categories*** |
|---|---|
| White British | White British |
| White Irish | White Irish |
| White Other | White Other |
| Mixed White-Black African | Mixed White-Black African |
| Mixed White-Black Caribbean | Mixed White-Black Caribbean |
| Mixed White-Asian | Mixed White-Asian |
| Mixed Other | Mixed Other |
| Black African | Black African |
| Black Caribbean | Black Caribbean |
| Black Other | Black Other |
| Indian | Indian |
| Pakistani | Pakistani |
| Bangladeshi | Bangladeshi |
| Asian Other | Asian Other & Chinese |
| Chinese | Romani |
| Romani | Irish Traveller |
| Irish Traveller | Other |
| Other | |

**Supplementary Figure 1: Data Flow Summary**

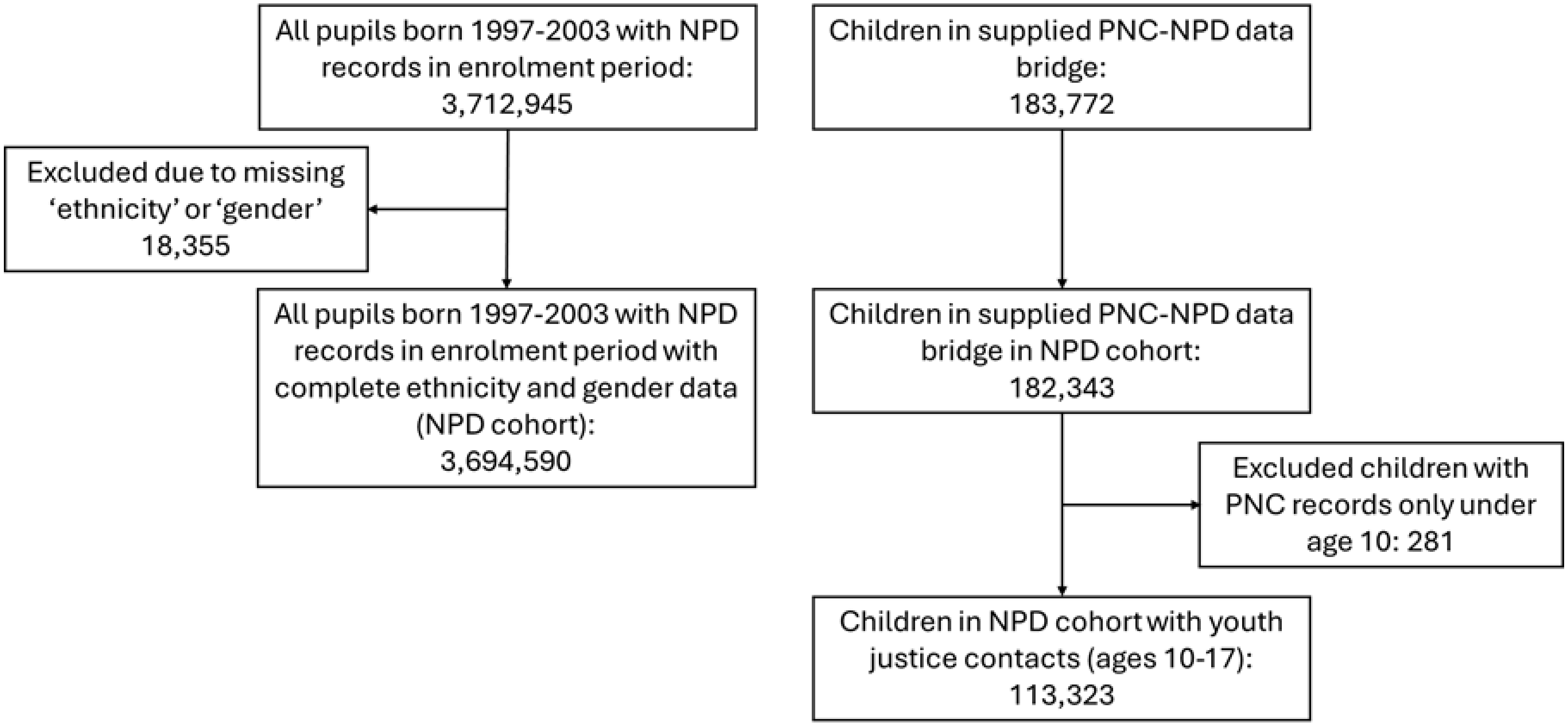

### Supplementary Table 2: Cohort Overview with Median Age at School Enrolment

| | White British | White Irish | White Other | Mixed White-Black African | Mixed White-Black Caribbean | Mixed White-Asian | Mixed Other | Black African | Black Caribbean | Black Other | Indian | Pakistani | Bangladeshi | Asian Other & Chinese | Romani | Irish Traveller | Other | Combined |
|---|---|---|---|---|---|---|---|---|---|---|---|---|---|---|---|---|---|---|
| **Pupils Included in the Study** ***n (%)*** | | | | | | | | | | | | | | | | | | |
| **All** | 2,629,227 | 13,627 | 211,850 | 18,755 | 46,768 | 35,192 | 59,126 | 130,356 | 52,212 | 23,294 | 101,792 | 146,947 | 59,131 | 77,564 | 12,978 | 3,970 | 71,801 | 3,694,590 |
| *(row%)* | *(71.2%)* | *(0.4%)* | *(5.7%)* | *(0.5%)* | *(1.3%)* | *(1.0%)* | *(1.6%)* | *(3.5%)* | *(1.4%)* | *(0.6%)* | *(2.8%)* | *(4.0%)* | *(1.6%)* | *(2.1%)* | *(0.4%)* | *(0.1%)* | *(1.9%)* | *(100.0%)* |
| **Male** | 1,348,304 | 6,938 | 108,263 | 9,366 | 23,682 | 17,975 | 30,253 | 66,079 | 26,434 | 11,895 | 52,793 | 75,687 | 30,205 | 40,236 | 6,649 | 2,056 | 37,684 | 1,894,499 |
| *(col%)* | *(51.3%)* | *(50.9%)* | *(51.1%)* | *(49.9%)* | *(50.6%)* | *(51.1%)* | *(51.2%)* | *(50.7%)* | *(50.6%)* | *(51.1%)* | *(51.9%)* | *(51.5%)* | *(51.1%)* | *(51.9%)* | *(51.2%)* | *(51.8%)* | *(52.5%)* | *(51.3%)* |
| **Female** | 1,280,923 | 6,689 | 103,587 | 9,389 | 23,086 | 17,217 | 28,873 | 64,277 | 25,778 | 11,399 | 48,999 | 71,260 | 28,926 | 37,328 | 6,329 | 1,914 | 34,117 | 1,800,091 |
| *(col%)* | *(48.7%)* | *(49.1%)* | *(48.9%)* | *(50.1%)* | *(49.4%)* | *(48.9%)* | *(48.8%)* | *(49.3%)* | *(49.4%)* | *(48.9%)* | *(48.1%)* | *(48.5%)* | *(48.9%)* | *(48.1%)* | *(48.8%)* | *(48.2%)* | *(47.5%)* | *(48.7%)* |
| **Median Age at School Enrolment** ***(years)*** | | | | | | | | | | | | | | | | | | |
| **All** | 5.20 | 5.21 | 5.88 | 5.21 | 5.20 | 5.21 | 5.21 | 5.21 | 5.21 | 5.21 | 5.21 | 5.21 | 5.21 | 5.30 | 6.30 | 5.21 | 5.30 | 5.21 |
| **Male** | 5.20 | 5.21 | 5.79 | 5.21 | 5.20 | 5.21 | 5.21 | 5.21 | 5.21 | 5.21 | 5.21 | 5.21 | 5.21 | 5.30 | 6.38 | 5.21 | 5.30 | 5.21 |
| **Female** | 5.20 | 5.21 | 5.88 | 5.21 | 5.20 | 5.21 | 5.21 | 5.21 | 5.21 | 5.21 | 5.21 | 5.21 | 5.21 | 5.30 | 6.21 | 5.21 | 5.30 | 5.21 |

### Supplementary Table 3: Rates per 1,000 children and young people

| | White British | White Irish | White Other | Mixed White-Black African | Mixed White-Black Caribbean | Mixed White-Asian | Mixed Other | Black African | Black Caribbean | Black Other | Indian | Pakistani | Bangladeshi | Asian Other & Chinese | Romani | Irish Traveller | Other | Combined |
|---|---|---|---|---|---|---|---|---|---|---|---|---|---|---|---|---|---|---|
| **(a) Alternative Provision** ***Rates per 1,000 (95% CI)*** | | | | | | | | | | | | | | | | | | |
| **All** | 11.7 (11.6-11.8) | 16.1 (14.0-18.3) | 10.5 (10.0-10.9) | 17.0 (15.1-18.8) | 28.5 (27.0-30.0) | 10.6 (9.5-11.7) | 16.9 (15.8-17.9) | 12.6 (12.0-13.2) | 29.9 (28.5-31.4) | 20.2 (18.4-22.0) | 4.7 (4.3-5.1) | 7.1 (6.7-7.5) | 5.8 (5.2-6.4) | 7.3 (6.7-7.9) | 60.6 (56.5-64.7) | 86.9 (78.1-95.7) | 14.0 (13.1-14.8) | 12.0 (11.9-12.2) |
| **Male** | 16.8 (16.6-17.0) | 22.5 (19.0-26.0) | 13.5 (12.8-14.1) | 23.5 (20.4-26.6) | 40.7 (38.2-43.3) | 14.7 (13.0-16.5) | 23.4 (21.7-25.1) | 17.8 (16.8-18.8) | 45.2 (42.7-47.7) | 29.3 (26.3-32.4) | 5.8 (5.2-6.5) | 9.8 (9.1-10.5) | 8.0 (7.0-9.0) | 9.8 (8.8-10.7) | 69.6 (63.5-75.8) | 105.1 (91.8-118.3) | 18.8 (17.4-20.2) | 17.0 (16.8-17.2) |
| **Female** | 6.4 (6.2-6.5) | 9.6 (7.2-11.9) | 7.4 (6.8-7.9) | 10.4 (8.4-12.5) | 15.9 (14.2-17.5) | 6.3 (5.1-7.5) | 10.0 (8.8-11.1) | 7.3 (6.7-8.0) | 14.3 (12.9-15.8) | 10.6 (8.7-12.5) | 3.5 (2.9-4.0) | 4.3 (3.8-4.7) | 3.5 (2.8-4.2) | 4.7 (4.0-5.4) | 51.2 (45.8-56.6) | 67.4 (56.2-78.6) | 8.6 (7.7-9.6) | 6.8 (6.7-7.0) |
| **(b) Alternative Provision among Pupils Ever Recorded with SEND-for-SEMH** ***Rates per 1,000 (95% CI)*** | | | | | | | | | | | | | | | | | | |
| **All** | 80.8 (79.6-81.9) | 115.6 (95.7-135.6) | 58.9 (54.2-63.6) | 94.8 (81.5-108.2) | 127.1 (119.5-134.8) | 72.8 (62.3-83.3) | 95.1 (87.2-103.0) | 59.3 (54.7-63.9) | 113.5 (106.9-120.0) | 94.4 (82.9-105.9) | 24.7 (18.7-30.7) | 44.9 (40.0-49.7) | 37.8 (30.7-44.9) | 38.7 (30.3-47.1) | 134.5 (115.8-153.3) | 191.9 (159.5-224.3) | 49.8 (42.7-56.9) | 79.9 (79.0-80.9) |
| **Male** | 91.9 (90.5-93.4) | 137.1 (111.1-163.1) | 67.0 (61.0-73.0) | 102.3 (85.8-118.9) | 145.5 (135.7-155.4) | 85.7 (71.9-99.5) | 107.1 (97.0-117.3) | 67.9 (62.1-73.7) | 130.1 (121.8-138.5) | 110.9 (96.3-125.6) | x | 52.2 (46.1-58.4) | 47.0 (37.3-56.8) | x | 151.4 (128.0-174.9) | 204.5 (165.0-244.0) | 57.6 (48.5-66.7) | 91.2 (89.9-92.4) |
| **Female** | 55.5 (53.7-57.2) | 69.8 (41.7-98.0) | 41.4 (34.4-48.5) | 77.7 (55.7-99.8) | 87.8 (76.2-99.4) | 46.0 (31.1-60.9) | 69.6 (57.4-81.8) | 38.3 (31.4-45.3) | 75.5 (65.7-85.3) | 53.9 (37.5-70.4) | x | 27.0 (20.0-34.0) | 20.7 (11.7-29.6) | x | 93.8 (64.2-123.4) | 161.7 (105.8-217.5) | 31.9 (21.5-42.3) | 54.6 (53.1-56.1) |
| **(c) Alternative Provision among Pupils Ever Recorded with Youth Justice Contact** ***Rates per 1,000 (95% CI)*** | | | | | | | | | | | | | | | | | | |
| **All** | 127.0 (124.7-129.3) | 145.1 (114.5-175.7) | 81.4 (73.5-89.3) | 131.7 (109.8-153.5) | 161.5 (149.9-173.1) | 109.4 (89.9-128.9) | 147.2 (132.9-161.5) | 87.1 (78.8-95.5) | 174.6 (163.0-186.2) | 138.6 (119.3-157.9) | 46.1 (30.5-61.7) | 68.3 (59.7-76.9) | 57.1 (45.0-69.3) | 64.9 (47.1-82.6) | 141.0 (122.8-159.2) | 151.8 (125.3-178.3) | 85.4 (71.8-99.0) | 123.3 (121.4-125.2) |
| **Male** | 133.3 (130.6-135.9) | 146.8 (111.9-181.7) | 82.4 (73.6-91.2) | 123.1 (99.4-146.8) | 170.7 (157.1-184.2) | 112.2 (90.3-134.2) | 147.0 (131.0-163.0) | 84.2 (75.5-93.0) | 176.1 (163.4-188.8) | 137.1 (116.5-157.7) | x | 69.5 (60.5-78.6) | x | x | 146.2 (124.6-167.8) | 161.9 (130.4-193.4) | 82.9 (68.7-97.2) | 127.2 (125.0-129.3) |
| **Female** | 105.8 (101.4-110.3) | 139.1 (75.9-202.4) | 77.4 (59.6-95.2) | 166.7 (112.2-221.1) | 130.8 (108.6-153.0) | 97.9 (56.1-139.8) | 147.8 (116.3-179.4) | 107.8 (81.6-134.0) | 166.9 (138.8-195.0) | 148.1 (93.4-202.9) | x | 53.8 (26.4-81.3) | x | x | 127.0 (93.4-160.5) | 122.2 (74.4-170.1) | 105.0 (60.3-149.6) | 108.7 (104.8-112.7) |

**Supplementary Table 4: Median Age-at-First (a) Alternative Provision enrolment among all school-aged children (b) Alternative Provision enrolment among pupils ever recorded with SEND-for-SEMH (c) Alternative Provision enrolment among pupils ever recorded with Youth Justice Contact (d) SEND-for-SEMH among pupils ever recorded with Alternative Provision (e) Youth Justice Contact among pupils ever recorded Alternative Provision**

| | White British | White Irish | White Other | Mixed White-Black African | Mixed White-Black Caribbean | Mixed White-Asian | Mixed Other | Black African | Black Caribbean | Black Other | Indian | Pakistani | Bangladeshi | Asian Other & Chinese | Romani | Irish Traveller | Other | Combined |
|---|---|---|---|---|---|---|---|---|---|---|---|---|---|---|---|---|---|---|
| **(a) Age at First Alternative Provision Enrolment: All school-aged Children, *Median Age (years)*** | | | | | | | | | | | | | | | | | | |
| **All** | 13.8 | 14.1 | 14.9 | 13.9 | 14.0 | 13.7 | 13.9 | 14.2 | 13.7 | 14.4 | 14.8 | 14.6 | 14.6 | 15.0 | 13.8 | 12.6 | 15.4 | 13.9 |
| **Male** | 13.4 | 13.5 | 14.7 | 13.4 | 13.7 | 13.5 | 13.4 | 14.1 | 13.4 | 14.2 | 14.7 | 14.3 | 14.3 | 15.0 | 13.8 | 12.6 | 15.3 | 13.6 |
| **Female** | 14.5 | 14.6 | 15.3 | 14.7 | 14.8 | 14.8 | 14.6 | 14.6 | 14.4 | 14.6 | 15.2 | 15.0 | 15.3 | 15.4 | 13.7 | 12.6 | 15.6 | 14.6 |
| **(b) Age at First Alternative Provision Enrolment: Pupils ever recorded with SEND-for-SEMH, Median Age (years)** | | | | | | | | | | | | | | | | | | |
| **All** | 13.7 | 13.0 | 14.1 | 13.8 | 13.8 | 14.1 | 13.6 | 13.4 | 13.7 | 13.7 | 14.8 | 14.3 | 14.2 | 14.2 | 13.6 | 13.2 | 14.5 | 13.8 |
| **Male** | 13.5 | 12.9 | 13.9 | 13.4 | 13.6 | 13.9 | 13.3 | 13.3 | 13.5 | 13.7 | x | 14.2 | 14.1 | x | 13.4 | 13.0 | 14.3 | 13.5 |
| **Female** | 14.4 | 14.0 | 14.7 | 14.2 | 14.7 | 14.5 | 14.4 | 13.9 | 14.3 | 13.9 | x | 14.8 | 14.7 | x | 14.0 | 13.4 | 15.4 | 14.4 |
| **(c) Age at First Alternative Provision Enrolment: Pupils ever recorded with Youth Justice Contact, Median Age (years)** | | | | | | | | | | | | | | | | | | |
| **All** | 14.1 | 14.0 | 14.8 | 14.2 | 14.2 | 14.3 | 14.3 | 14.4 | 14.1 | 14.4 | 15.1 | 14.8 | 14.6 | 14.8 | 13.7 | 13.7 | 15.0 | 14.2 |
| **Male** | 14.0 | x | 14.7 | 14.1 | 14.0 | x | 14.2 | 14.4 | 14.0 | 14.4 | x | x | x | x | 13.7 | 13.8 | x | 14.1 |
| **Female** | 14.5 | x | 14.9 | 15.0 | 14.6 | x | 14.5 | 14.5 | 14.4 | 14.0 | x | x | x | x | 14.0 | 13.5 | x | 14.5 |
| **(d) Age at First SEND-for-SEMH: Pupils ever recorded with Alternative Provision, Median Age (years)** | | | | | | | | | | | | | | | | | | |
| **All** | 9.2 | 9.3 | 11.2 | 9.3 | 8.8 | 10.9 | 8.8 | 8.9 | 8.2 | 8.3 | 11.5 | 11.3 | 12.7 | 11.4 | 11.4 | 9.3 | 11.2 | 9.3 |
| **Male** | 8.8 | 8.5 | 10.4 | 8.1 | 8.2 | 10.6 | 7.9 | 8.4 | 7.8 | 8.3 | x | 11.2 | 12.4 | x | 11.4 | 8.5 | 9.8 | 8.8 |
| **Female** | 11.7 | 10.9 | 12.8 | 12.3 | 11.6 | 11.7 | 11.9 | 12.2 | 10.5 | 8.5 | x | 12.0 | 13.8 | x | 11.6 | 11.2 | 12.6 | 11.7 |
| **(e) Age at First Youth Justice Contact: Pupils ever recorded Alternative Provision, Median Age (years)** | | | | | | | | | | | | | | | | | | |
| **All** | 13.9 | 13.8 | 14.5 | 13.8 | 14.0 | 14.4 | 14.2 | 14.4 | 14.3 | 14.4 | 14.7 | 14.6 | 14.5 | 14.6 | 14.1 | 13.8 | 14.8 | 14.0 |
| **Male** | 13.8 | x | 14.5 | 13.7 | 14.0 | x | 14.2 | 14.5 | 14.3 | 14.5 | x | x | x | x | 13.9 | 13.9 | x | 14.0 |
| **Female** | 14.1 | x | 14.5 | 14.0 | 14.2 | x | 14.3 | 14.4 | 14.2 | 14.3 | x | x | x | x | 14.4 | 13.4 | x | 14.1 |
